# Creating a verified health facility dataset for spatial analysis: example from Greater Accra, Ghana

**DOI:** 10.64898/2026.09.30.26364381

**Authors:** Elizabeth J. Mroz, Erica Afumaa, Amanda J. Mason-Jones

## Abstract

Geocoded health facility data are essential for spatial analyses, yet routine health information systems in many low- and middle-income countries often contain inaccurate, incomplete, inconsistent, or outdated facility coordinates. For Greater Accra, Ghana, we created a geocoded health facility dataset aligned with routine maternal health service data from the District Health Information Management System (DHIMS2). We undertook a desk-based, multi-source geocoding and verification workflow of 602 facilities reporting maternal health services between 2020 and 2024. Candidate coordinates were triangulated using national facility records, online mapping platforms, facility websites and social media, directories, photographs, satellite imagery, and local expert knowledge. Coordinates were classified according to the strength and concordance of supporting evidence, with uncertain cases undergoing secondary review. Coordinates were identified for 96.4% of 602 health facilities (*n* = 580), with 59.3% assigned a high confidence classification (*n* = 357). Only 22 facilities (3.7%) could not be located with sufficient supporting evidence. Our resulting dataset is made available in a version-controlled openly-modifiable repository for long-term use, permitting routine service utilisation data to be linked with facility locations; thereby facilitating spatial analysis important for equitable health service planning. By documenting our process, this study makes a typically hidden stage of spatial health research transparent and reproducible. Our findings also highlight the need to treat geocoded facility datasets as living, maintainable geospatial health data infrastructure rather than as static research outputs.

## Introduction

Geocoded health facility data are essential for geographical accessibility modelling and spatial analyses of health services (1,2). Yet, in low- and middle-income countries, national facility lists may be absent, imprecise, or no longer correspond to the current facility location (1,3–5).These problems are particularly important where facilities open, close, relocate, change name, or change function over time. As accurate facility locations are crucial inputs to travel-time and other spatial health analyses, positional errors resulting from poor spatial and temporal alignment can substantially alter estimates of healthcare accessibility and spatial equity (4).

Considerable effort has previously been made to provide geocoded health facility information in sub-Saharan Africa where not reliably available. Regional, country-specific, and even continent-wide reconciled health facility datasets have been assembled (1,4,5). However, an important methodological and infrastructural problem remains. Such datasets may be compiled as part of a wider study, with the resulting work not published; this subsequently results in duplication of effort where others wish to undertake spatial analysis but cannot access this data (2). Even where considerable effort has been undertake to generate large datasets, they are often static snapshots rather than actively-maintained systems (2,3,5). This places a critical limitation on the longevity of these datasets, as health systems are dynamic with facilities coverage commonly changing. Consequently, recent work has emphasised the need for geospatial facility data to be treated as living health-system infrastructure with regular updating, clear stewardship, and open access, rather than continuing the pattern of constructing these datasets as static one-off research outputs (2,6)

Ghana provides a useful case study to examine this specific problem. Ghana’s District Health Information Management System (DHIMS2) is the national repository for reporting routine health service data (7), including maternal and neonatal health indicators (8,9). The Ghana Health Service (GHS) separately maintains a directory of health facility locations on its public-facing website. However, in our initial comparison, the GHS and DHIMS2 records were not directly aligned, creating discrepancies in facility names, coverage, status, and crucially, geographic coordinates. Ghana is a highly relevant setting for spatial accessibility research, given existing work examining geographic inequalities in access to health services and their implications for health planning (10,11). Ghana’s Ministry of Health continues to remain committed to achieving universal health coverage, and has flagged geographic accessibility modelling as a key methodology for evaluating health equity and informing health service planning (7,12–14).

Our need for a verified health facility dataset for Ghana emerged during a wider project examining maternal healthcare access under flooding and future climate-change scenarios. We intended to undertake geospatial accessibility modelling, and obtained monthly aggregated maternal health service utilisation data for Greater Accra from DHIMS2. However, the provided dataset did not include facility coordinates. We therefore attempted to link the DHIMS2 facility records to coordinates available through GHS, however the linkage was not reliable due to mismatched facility names, facilities appearing in DHIMS2 but not GHS, duplicate records in GHS, obviously incorrect or approximated coordinates, and evidence that some facilities had possibly closed or relocated without corresponding updates being made in DHIMS2. Neither the DHIMS2 nor the GHS data were provided with temporal metadata with which to assess when such changes had occurred. These inconsistencies prevented the direct use of the routine service-utilisation data in the planned spatial analyses.

Similar problems have been documented in health facility inventories across sub-Saharan Africa, including discrepancies between facility lists, missing or conflicting coordinates, uncertain operational status, and limited mechanisms for updating records (1–4). In a review of Ghana’s DHIMS2 conducted in August 2025, DHIMS2 recorded 11,129 health facilities nationally, of which 1,752 were sampled for assessment; of these, 101 sampled facilities were reported to be non-functional (6%) (7). In Greater Accra, our study area, 31 of 168 sampled facilities were reported to be non-functional (18%) (7).

Despite the frequency of these problems, the practical process used to reconcile and obtained reliable coordinates is often reported only briefly and in passing. Terms such as “*manual delineation”* can obscure the substantial work involved in resolving mismatched names, duplicate records, conflicting coordinates, changing facility status, and uncertainty about specific locations (1,4). Additionally, the limited documentation of the data curation process continues to be largely invisible in published studies, resulting in limited methodological understanding of these decisions; this reduces reproducibility and makes it more likely researchers will repeatedly construct facility datasets from the beginning rather than reuse and progressively improve resources (2,5). Transparent methodological documentation is therefore needed not only to show how locations were obtained, but also to demonstrate how conflicting evidence was evaluated and how uncertainty was retained and communicated.

We therefore used the data-alignment problem encountered in Greater Accra as an opportunity to make this otherwise hidden curation process explicit. We aimed to construct a geocoded health facility dataset aligned with routine maternal health service data for Greater Accra, Ghana, while documenting a transparent, multi-source workflow for resolving conflicting or incomplete facility information. Rather than developing a novel geocoding algorithm, our objective was to describe the evidential and decision-making process required to reconcile routine facility records and to publish the resulting dataset in a version-controlled repository so that it could be inspected, reused, corrected, and progressively updated. In doing so, the study addresses both an immediate analytical barrier and a broader challenge in geospatial health-data infrastructure: how facility datasets can be treated as maintainable resources rather than one-off research outputs.

## Methods

### Study area

Greater Accra is a region in Ghana, with a dense metropolitan city (Accra) surrounded by peri-urban and rural districts (15). In DHIMS2, the region contains 602 health facilities which report offering maternal health services between 2020 and 2024. These facilities span primary, secondary, and tertiary care offered by public, private, and faith-based providers, with facility types including hospitals, health centres, and CHPS (Community-based Health Planning and Services, a national strategy designed to bring basic healthcare to rural communities) (16).

### Study design and data source

This retrospective, desk-based study was undertaken within a wider project investigating maternal healthcare access under flooding and future climate-change scenarios in Greater Accra. Monthly aggregated maternal health-service utilisation data were obtained from DHIMS2 for January 2020 through December 2024. DHIMS2 is Ghana’s web-based system for collating and reporting routine health-service data (8,9). The supplied records identified 602 facilities but did not contain geographic coordinates; these records therefore formed the facility set for the geocoding exercise. The objective was to assign a defensible location to each DHIMS2 facility record while preserving the facility names used in DHIMS2 so that coordinates could subsequently be linked to the routine utilisation data. In this study, verification refers to desk-based corroboration of facility identity and location across multiple evidence streams. It does not imply field-based validation or GPS ground-truthing.

### Identification of possible facility locations

Each DHIMS2 facility record underwent manual review using the facility name and available administrative information as the starting point (Figure 1). Candidate locations were sought across multiple sources, including the Ghana Health Service facility directory and national facility records; online mapping platforms including Google Maps, OpenStreetMap, Yandex Maps and Mapcarta; non-governmental and health-sector directories; facility websites; publicly accessible social-media pages; news reports; local directories; public reviews and photographs; satellite imagery; and local expert knowledge. No source was treated as authoritative a priori. Instead, candidate locations were assessed using distinct evidence streams that differed in their likely provenance, specificity, currency, and spatial precision. Existing coordinates were therefore treated as candidate locations rather than accepted automatically

**Figure 1.**
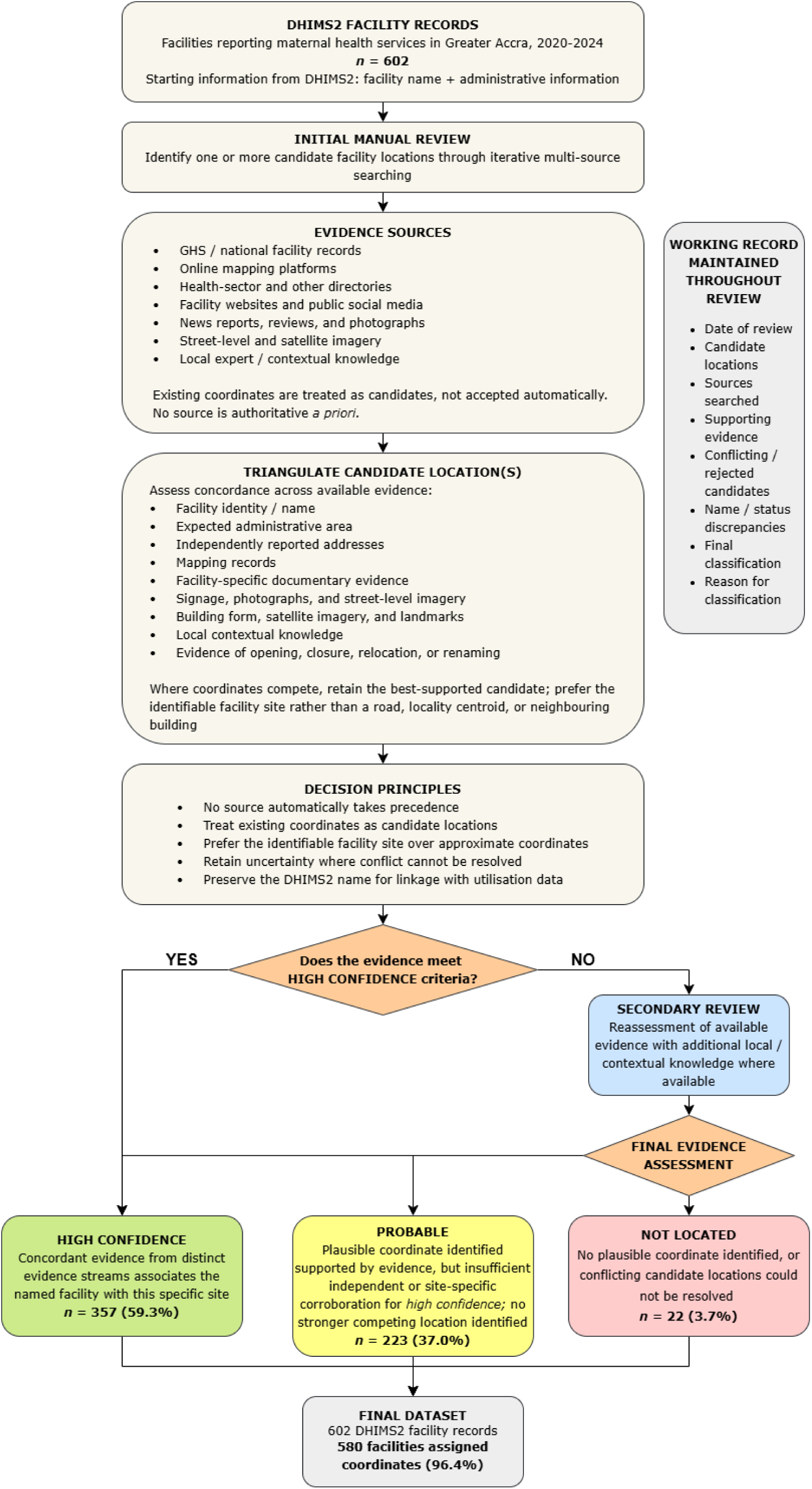
Workflow for desk-based geocoding and verification of DHIMS2 health facility records in Greater Accra, Ghana.

Searches were iterative rather than conducted according to a fixed hierarchy of sources. Where an exact facility name did not identify a plausible location, searches incorporated spelling variants, alternative or previous facility names, relevant districts or sub-districts, facility type and other contextual information where available. Where the same or similar facility name occurred at multiple locations, these were investigated as potentially distinct facilities or branches rather than automatically treated as duplicates.

### Triangulation of coordinates

Candidate locations were assessed against the facility identity and administrative information recorded in DHIMS2 and triangulated using the other available evidence. Agreement between facility name and expected administrative area was considered evidence that a candidate location was plausible, but was not sufficient on its own for a high-confidence classification. The multi-source reconciliation approach was informed by previous health-facility inventory studies that combined administrative lists, online mapping resources, imagery, manual review, and coordinate validation (1,3,4).

Where multiple candidate coordinates referred to the same facility and differed only at a local scale, preference was given to the coordinate judged to most closely represent the identifiable facility site or building. Coordinates falling on roads, approximate locality centroids or neighbouring buildings were not preferred solely because they originated from an official source. A GHS coordinate was preferentially retained only when it was consistent with the identified facility site and was of comparable or greater spatial precision than competing coordinates. GHS provenance alone was not sufficient to assign a high-confidence classification.

Where sources disagreed substantially, candidate coordinates were treated as competing locations. Evidence used to resolve disagreement included facility identity, administrative area, independently reported addresses, mapping records, facility websites and social-media information, photographs, street-level imagery, satellite imagery, surrounding landmarks and local contextual knowledge. No source automatically took precedence. Where the available evidence did not resolve disagreement sufficiently, uncertainty was retained in the final classification rather than assigning a high-confidence location.

### Visual evidence corroboration

Visual corroboration was considered present where street-level imagery or photographs showed facility signage, distinctive building characteristics, or surrounding landmarks that could be associated with a candidate site. Where possible, building form and surrounding features visible in photographs or street-level imagery were compared with satellite imagery. Satellite imagery alone was not considered sufficient evidence of facility identity unless it could be linked to facility-specific or contextual information.

### Confidence classification

Each facility was assigned one of three confidence classifications reflecting the strength and concordance of evidence supporting the assigned location (Table 1). These categories represent levels of evidential support for the association between a facility and a location and should not be interpreted as measures of positional accuracy.

**Table 1.** Operational definitions of facility location confidence classifications.

| Category | Operational definition |
| --- | --- |
| High confidence | A plausible coordinate with high confidence for a health facility, supported by concordant evidence from distinct evidence streams. Evidence is sufficient to associate this facility with this specific site, including agreement with either |
|  | independently-sourced address information, or visual evidence linking the named facility to this site. |
| Probable | A plausible coordinate supported by some evidence, including agreement in facility name and administrative area, but where independent corroboration between sources is insufficient or site-specific visual evidence is not available for a high-confidence classification. No stronger competing candidate location was identified. |
| Not located | No plausible coordinate could be identified. This is either due to insufficient evidence, or multiple conflicting candidate locations which cannot be resolved after all available sources have been reviewed. |

These classifications were based on evidential triangulation rather than a fixed numerical threshold for the number of supporting sources. Different sources were not considered equivalent: online mapping services and directories may reproduce information originating from the same underlying source, while publicly generated online content varies considerably in specificity, currency and reliability. Classification therefore reflected the specificity, concordance, independence and overall plausibility of the available evidence rather than a simple count of sources. The confidence classifications consequently group facilities with broadly similar levels of evidential support, while some variation in evidential strength remains within each category.

### Resolution of common discrepancies

A consistent set of decision principles was used to address recurring discrepancies encountered during review (Table 2). These principles formalised the main decision rules while allowing case-specific judgement where the available evidence differed substantially between facilities. They were not applied as an automatic algorithm.

**Table 2.**
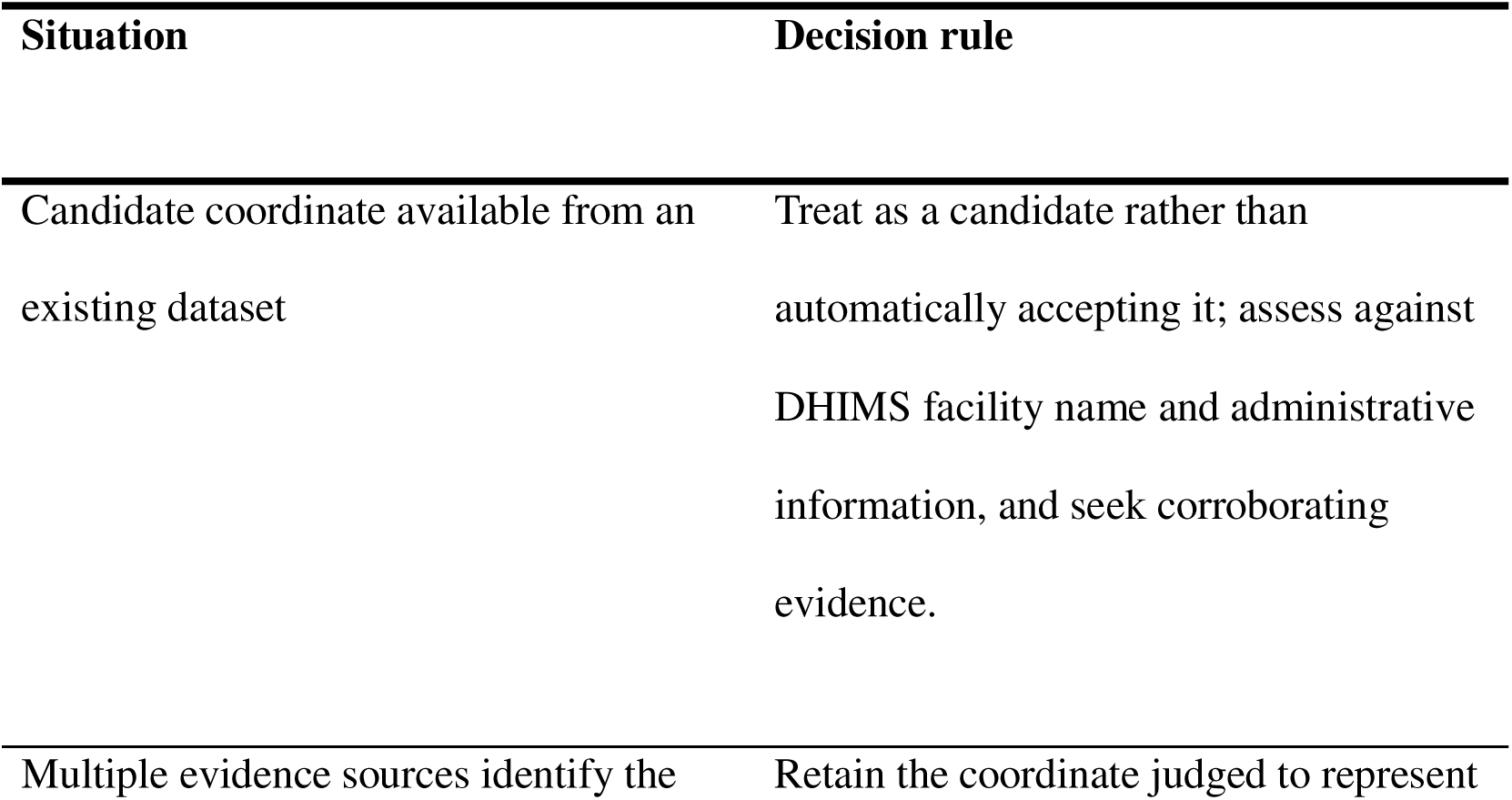
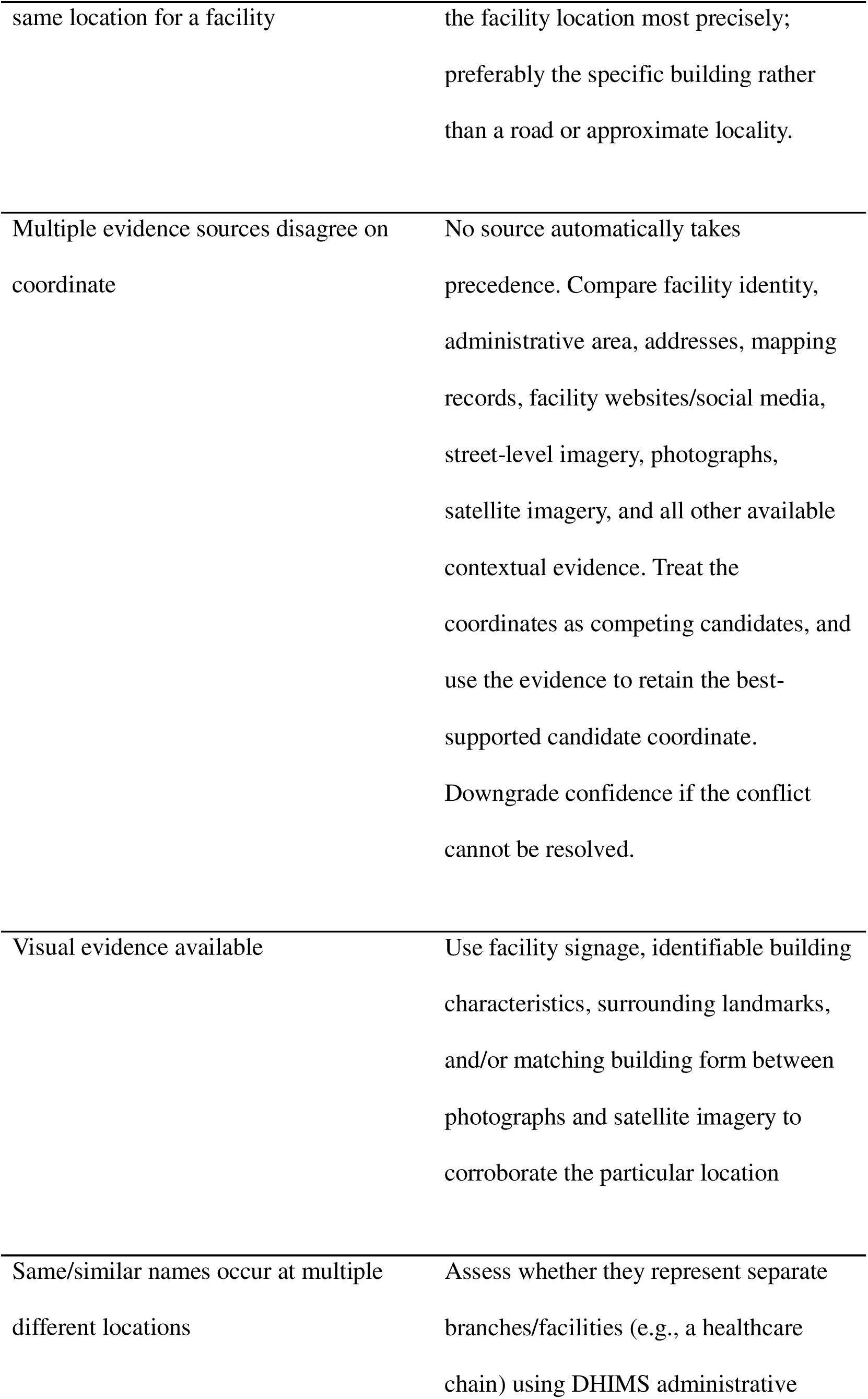
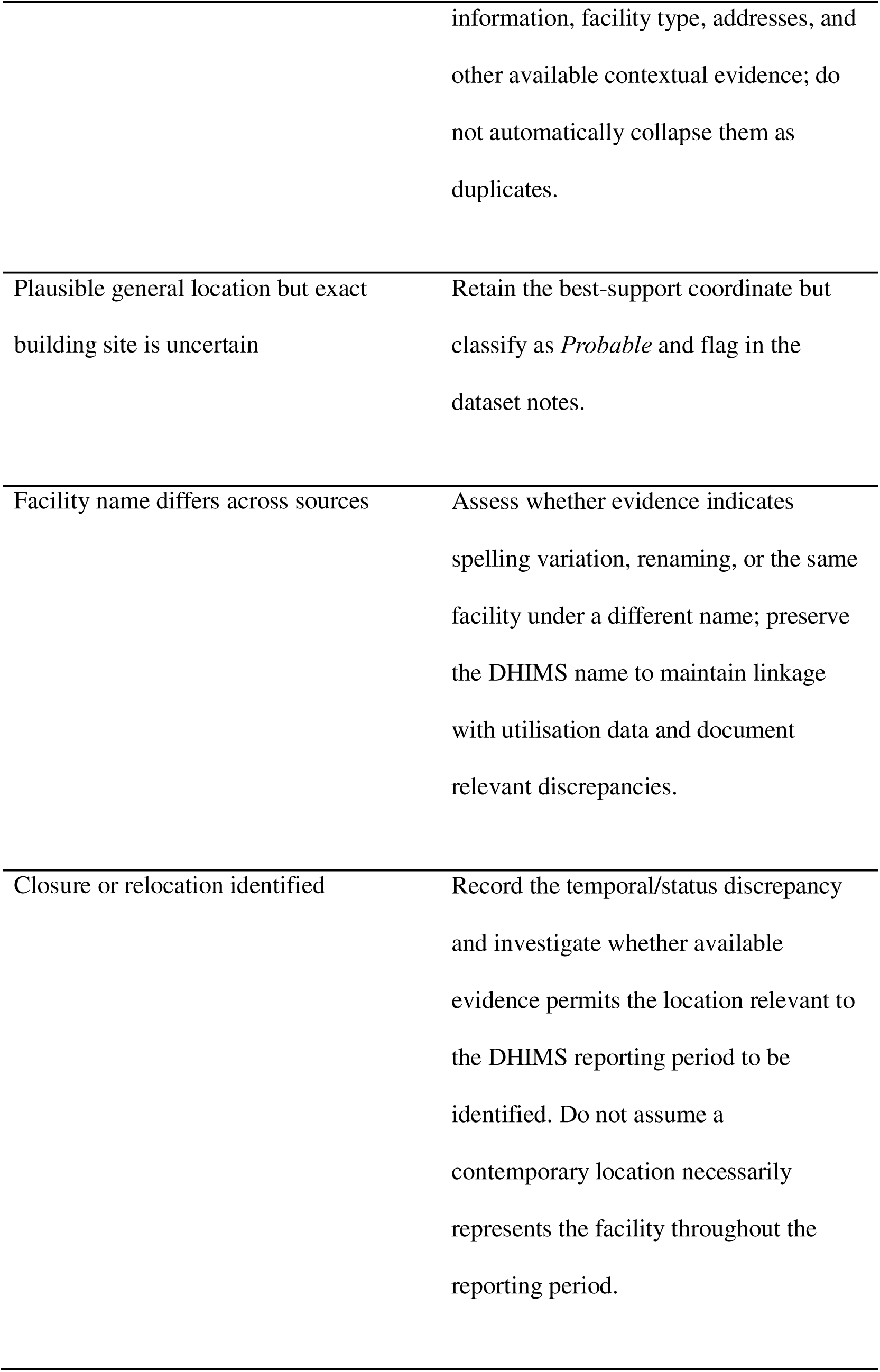
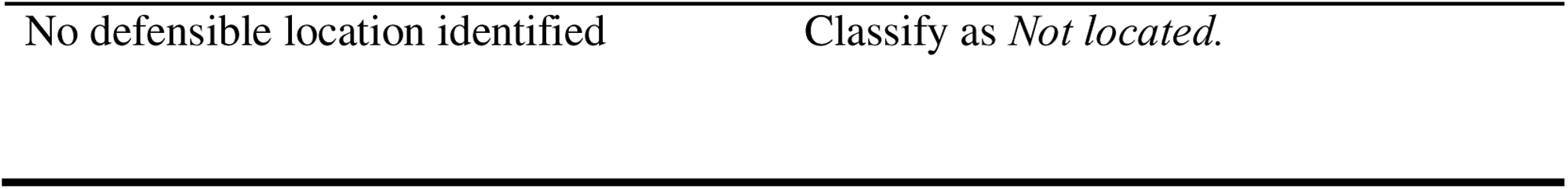
Set of decision principles used during facility geocoding and verification in order to resolve common discrepancies.

### Changes to facility information

Evidence suggesting that a facility had opened, closed, relocated, or changed name during or around the 2020–2024 reporting period was recorded during review. Where the available evidence permitted the location relevant to the DHIMS2 reporting period to be identified, this information was incorporated into the final location assignment. Where the timing or location of a change could not be established sufficiently, the discrepancy was retained in the working record and, where relevant, reflected in the confidence classification and public dataset notes. Contemporary online information was therefore not assumed to represent the facility throughout the complete reporting period.

### Initial review and secondary review process

All 602 facilities underwent an initial manual review by EM. Facilities for which the available evidence was insufficient to assign a High confidence classification underwent secondary review by EA, who reassessed the available evidence and could incorporate additional contextual knowledge of the study setting. Cases in which substantial uncertainty remained were discussed within the research team, with AMJ providing methodological oversight. Facilities meeting the High confidence criteria during the initial review were not routinely subjected to duplicate review.

Because secondary review was targeted towards facilities that could not be resolved confidently during the initial review, rather than being applied independently to the complete facility set or to a random sample, formal inter-reviewer agreement statistics were not calculated. Secondary review therefore functioned as an uncertainty-resolution and quality-control procedure rather than as a formal assessment of inter-reviewer reliability.

Additional quality-control procedures included maintaining a detailed working record of the evidence examined and the reasoning underlying each classification; escalating unresolved cases for secondary review; retaining uncertainty rather than forcing facilities into a High confidence category; recording coordinate provenance and confidence explicitly in the released dataset; and retaining relevant unresolved discrepancies in the dataset notes.

### Data recording, coordinate provenance, and maintenance of a working record

For each facility, the detailed working record documented the date of review, candidate locations, sources searched, source access dates where available, evidence supporting the selected location, conflicting or rejected candidates, relevant information concerning facility names or operational status, the final confidence classification, and, where applicable, the reason that a High confidence classification could not be assigned.

The publicly released dataset is a condensed output containing the retained coordinates, the source from which the final coordinate pair was obtained, the confidence classification, and additional notes where relevant uncertainty or discrepancies remained. The recorded coordinate source represents the provenance of the retained coordinate pair and should not be interpreted as the sole evidence used to verify the facility location. Assignment of a High confidence location depended on the wider triangulation process.

### Use of publicly-accessible digital information

Publicly accessible digital material was used only to corroborate facility-level characteristics, including facility identity, location, and operational status. Individuals represented in these sources were not study subjects. Researchers did not contact users or seek access to private or restricted content. Where user-generated material or individual professional profiles were consulted, only information necessary to resolve facility identity or location was extracted.

Personal names, usernames, and other individual identifiers were excluded from the analytical dataset and public repository. Individual-level material was not reproduced in the manuscript. The released dataset contains facility-level locations and associated metadata only and contains no patient-level or individual-level data.

The detailed working record was retained by the research team for quality-control purposes but was not publicly released because it contained links to and excerpts from individual-level online material that were unnecessary for reuse of the resulting facility dataset.

## Results and discussion

### Outcome of the creation of a new geocoded dataset

We reviewed all 602 DHIMS2 facility records reporting maternal health services in Greater Accra. A defensible coordinate was identified to 580 facilities (96.4%). Of these, 357 facilities (59.3% of the full dataset) were assigned a high-confidence classification and 223 (37.0%) were classified as probable locations. For the remaining 22 facilities (3.7%), no location could be identified with sufficient supporting evidence and these were classified as not located. We published the resulting dataset in a version-controlled public repository to support reuse, review, correction, and future updating (17).

The distinction between high-confidence and probable locations was important. The availability of a coordinate did not necessarily indicate that a facility had been located with equal evidential certainty. Some facilities could be associated with a specific site using concordant documentary or visual evidence, whereas others could only be placed plausibly within an expected locality. The confidence classifications should therefore be interpreted as levels of evidential support for the facility–location association, rather than as measures of positional accuracy.

### Evaluation of the reconciliation process

Substantial inconsistencies were encountered across the data sources reviewed, including mismatched facility names, duplicate records, competing coordinates, approximate or road-based locations, and evidence of relocation, closure or renaming. These problems are consistent with previously documented data-quality issues in master facility lists and geocoded facility inventories (1,4). No single source consistently provided the most reliable information, supporting the use of multi-source triangulation rather than reliance on a fixed source hierarchy (1,3,4).

Different evidence sources were useful for resolving different problems. Formal facility records were important for identifying candidate facilities, while facility websites, social media, public reviews, local contextual knowledge and imagery were often required to resolve uncertainty about facility identity or exact location. Ground-level photographs and street-level imagery were particularly useful where signage, building form or surrounding landmarks could be linked to satellite imagery (1,3,4).

We also observed that digital visibility was uneven. Larger urban facilities and private healthcare chains generally had more online evidence available, whereas rural and lower-tier facilities, particularly CHPS compounds, were more difficult to verify. This means that confidence may partly reflect the availability of digital evidence rather than only the underlying accuracy of the facility record. More generally, the availability and quality of geospatial health data is known to vary substantially between countries and subnational areas (2,5,18)

Temporal mismatch was another recurring issue. The DHIMS2 data covered January 2020 to December 2024, whereas maps, websites, imagery and other digital sources were updated at different times. Evidence of facility opening, closure, relocation and renaming demonstrated that contemporary facility locations cannot always be assumed to represent the full historical reporting period. Similar temporal and operational-status challenges have been reported in facility inventories elsewhere in sub-Saharan Africa (2–4).

### Implications for geospatial health data infrastructure

The need for substantial manual reconciliation illustrates why geocoded facility datasets should not be treated solely as static research outputs. Even a carefully constructed dataset will become outdated as facilities open, close, relocate or change function. Large regional or national inventories remain valuable, but without mechanisms for continued review and updating their usefulness will decline over time.

A more sustainable model may therefore be to embed transparent geocoding, provenance recording, confidence assessment and updating procedures within locally maintained health-information systems. Previous work has identified fragmented facility-data management, missing unique identifiers, and weak integration between facility registries and routine information systems as barriers to effective reuse (19). The version-controlled dataset produced here provides a mechanism for review and correction, but long-term maintenance should ultimately depend on local institutional ownership rather than repeated researcher-led reconstruction (2,6,20).

### Limitations

This study has several limitations. First, the facility set was defined by facilities reporting maternal health services in DHIMS2 rather than by an independent census. DHIMS2 participation may be incomplete, particularly among private providers; previous studies in Greater Accra have reported limited participation of private facilities in routine reporting systems (8,9); smaller, lower-tier or private providers may therefore be underrepresented. The resulting dataset should consequently be interpreted as a geocoded dataset aligned to DHIMS2 rather than a complete census of all maternal-health facilities in Greater Accra.

Second, the workflow depended substantially on digitally available information. Online mapping, facility websites, social media, photographs and street-level imagery were unevenly distributed across Greater Accra. Urban, higher-tier and larger private facilities generally had more corroborating evidence available, whereas rural and lower-tier facilities, particularly CHPS compounds, were more difficult to verify. Confidence classifications may therefore partly reflect differences in digital visibility rather than only differences in the underlying accuracy of facility records.

Third, the sources used for triangulation were not temporally uniform. DHIMS2 utilisation data covered January 2020 to December 2024, while maps, imagery, websites, social-media content and directories were created or updated at different times. Although evidence of facility opening, closure, relocation and renaming was documented where identified, a complete time-resolved facility history could not be reconstructed for every facility.

Fourth, verification was desk-based rather than field-based. We did not visit facilities or independently collect GPS coordinates, and the confidence classifications therefore represent the strength of triangulated evidence rather than measured positional accuracy. Some coordinate errors may remain, particularly for facilities with limited online presence, newly established facilities, facilities with similar names, and rural facilities.

The workflow was also resource-intensive and was developed in response to an unanticipated data-alignment problem within a wider research project rather than as a prospective methodological evaluation. It has been applied only in Greater Accra, and its transferability to settings with different levels of rurality, digital coverage, facility density and health-information infrastructure has not yet been empirically tested.

Finally, public release of facility geolocations raises data-governance considerations. Although the released dataset contains facility-level rather than patient-level information and largely consolidates information that is individually discoverable through public sources, compilation into a single dataset reduces the effort required to identify facilities at scale. Decisions regarding publication of precise facility locations should therefore remain context-specific, particularly in insecure settings or where particular facility types may be sensitive.

### Future work

The immediate next application of the dataset will be spatial accessibility analyses examining women’s access to maternal health services in Greater Accra, including analyses under flood and future climate scenarios and sensitivity analyses comparing alternative geocoded facility datasets. This will allow the influence of facility-location data on subsequent accessibility estimates to be examined directly.

Formal review and integration by the Ghana Ministry of Health and Ghana Health Service would provide an important next step. Government-led review could provide additional confirmation of uncertain facility locations while supporting longer-term alignment between routinely collected service-utilisation data and geospatial facility information. Government stewardship and routine updating are widely identified as important for preventing facility datasets from becoming static (2,6,20). Particular attention may be required for rural and lower-tier facilities, which were more difficult to verify using desk-based sources.

The workflow should also be applied and evaluated in other settings. Although similar geocoding problems have been reported elsewhere in sub-Saharan Africa (1,3,4), this study was conducted only in Greater Accra and transferability has not yet been empirically demonstrated. Application in settings with different levels of rurality, digital coverage, facility density and health-information infrastructure would help establish which components of the workflow are broadly transferable and which require local adaptation.

More broadly, future work should consider how geocoding, verification and updating procedures can be incorporated into locally maintained health-information systems rather than relying on repeated researcher-led reconstruction of facility datasets. Common approaches to documenting provenance, confidence and version history could support interoperable datasets that can be corrected and progressively maintained as health systems change.

## Conclusion

A multi-source desk-based workflow enabled routine maternal health service data in Greater Accra to be linked to defensible facility locations while retaining uncertainty where evidence was insufficient. The study also makes explicit a data-curation process that is often hidden within spatial health research. More sustainable geospatial health-data infrastructure will require not only better facility inventories, but transparent and locally maintainable mechanisms for updating, provenance and review.

## Acknowledgements

Samuel Adjorlolo, Caroline Badzi, and Sly Moore are thanked for their assistance in procuring the DHIMS2 data. We thank the anonymous reviewer of an earlier version of this paper for a thoughtful evaluation of our work and constructive comments, which substantially strengthened the manuscript.

## Data availability

The minimal data set underlying the findings reported in this study is publicly available in Zenodo at https://doi.org/10.5281/zenodo.22877900. An actively maintained, version-controlled version of the dataset is also available through the project GitHub repository at https://github.com/FloodMums/greater-accra-health-facilities

## Funding

This work was funded by the Medical Research Foundation Climate Change and Health programme (MRF-RGL-ICCH-23-155) for the project *The impact of flood events on maternal health in two coastal cities: Accra and Kingston upon Hull*. AMJ was the PI of this award, and EJM was a Co-I. The Medical Research Foundation (hwww.medicalresearchfoundation.org.uk/) did not play any role in the study design, data collection and analysis, decision to publish, or preparation of the manuscript.

## Competing interests

The authors have declared that no competing interests exist.

## Ethics statement

Formal written consent of ethical approval was obtained from the Ghana Health Service Ethics Review Committee, of which use of DHIMS2 data was approved GHS-ERC: 022/10/24).

## Notes

### Competing Interest Statement

The authors have declared no competing interest.

## References

1. Maina J, Ouma PO, Macharia PM, Alegana VA, Mitto B, Fall IS, et al. A spatial database of health facilities managed by the public health sector in sub Saharan Africa. Sci Data. 2019 Jul 25;6(1):134. doi:10.1038/s41597-019-0142-2

2. Macharia PM, Beňová L, Ray N, Semaan A, Musau MM, Kipterer JK, et al. Putting health facilities on the map: a renewed call to create geolocated, comprehensive, updated, openly licensed dataset of health facilities in sub-Saharan African countries. BMC Med. 2025 Apr 7;23(1):211. doi:10.1186/s12916-025-04023-z

3. Moturi AK, Suiyanka L, Mumo E, Snow RW, Okiro EA, Macharia PM. Geographic accessibility to public and private health facilities in Kenya in 2021: An updated geocoded inventory and spatial analysis. Front Public Health. 2022 Nov 3;10. doi:10.3389/fpubh.2022.1002975

4. Gueye DM, Ly AB, Gueye B, Ndour PI, Fullman N, Liu PY, et al. A consolidated and geolocated facility list in Senegal from triangulating secondary data. Sci Data. 2024 Jan 24;11(1):119. doi:10.1038/s41597-024-02968-z

5. South A, Dicko A, Herringer M, Macharia PM, Maina J, Okiro EA, et al. A reproducible picture of open access health facility data in Africa and R tools to support improvement. Wellcome Open Res. 2021 Feb 16;5:157. doi:10.12688/wellcomeopenres.16075.2 PubMed PMID: 33437875; PubMed Central PMCID: PMC7780339.

6. Nshuti MAB, Touray K, Muluh TJ, Ubong GA, Ngofa RO, Mohammed BI, et al. Development of a Consolidated Health Facility Masterlist Using Data From Polio Electronic Surveillance in the World Health Organization African Region. JMIR Public Health and Surveillance. 2024 Jun 21;10(1):e54250. doi:10.2196/54250

7. Ghana Health Service. Ghana Harmonised Health Facility Assessment, Quality of Care Survey, 2025 [Internet]. Accra: Ghana Health Service; 2025.

8. Amoakoh-Coleman M, Kayode GA, Brown-Davies C, Agyepong IA, Grobbee DE, Klipstein-Grobusch K, et al. Completeness and accuracy of data transfer of routine maternal health services data in the greater Accra region. BMC Res Notes. 2015 Apr 1;8(1):114. doi:10.1186/s13104-015-1058-3

9. Kayode GA, Amoakoh-Coleman M, Brown-Davies C, Grobbee DE, Agyepong IA, Ansah E, et al. Quantifying the Validity of Routine Neonatal Healthcare Data in the Greater Accra Region, Ghana. PLOS ONE. 2014 Aug 21;9(8):e104053. doi:10.1371/journal.pone.0104053

10. Korah PI, Nunbogu AM, Ahmed A. Measuring access to health facilities in Ghana: Implications for implementation of health interventions and the Sustainable Development Goal 3. Applied Geography. 2023 Sep 1;158:103026. doi:10.1016/j.apgeog.2023.103026

11. Dumedah G, Iddrisu S, Asare C, Adu-Prah S, English S. Inequities in spatial access to health services in Ghanaian cities. Health Policy Plan. 2023 Dec 1;38(10):1166–80. doi:10.1093/heapol/czad084

12. Ochoa, CR. Geographic accessibility to equitable, basic and essential healths ervices in Ghana. Main technical report (April 2025). Accra: Ghana Health Service; 2025.

13. Ministry of Health, Ghana. Ghana’s Roadmap for Universal Health Coverage (2020-2030) [Internet]. Accra: Ministry of Health; 2020.

14. World Health Organisation. Geographic accessibility to health services in Ghana: Technical Report (May 2024). Geneva: World Health Organisation; 2024.

15. Siabi EK, Kabo-bah AT, Akpoti K, Anornu GK, Mortey EM, Awafo EA, et al. Rising urban flood risk and exposure under shared socioeconomic pathways in Greater Accra Ghana. Sci Rep. 2026 Apr 10;16(1):16105. doi:10.1038/s41598-026-47979-5

16. Asuo-Mante E, Mumuni AJ, Asuo-Mante M, Yelifari L, Awoonor-Williams JK, Phillips JF. A Pilot Trial of Applying Geographic Information System Technology to Health System Strengthening in the Upper East Region of Ghana. JMID. 2017 Nov 16;1(1):1–9. doi:10.14302/issn.2641-5526.jmid-17-1762

17. Mroz E, Afumaa E, Mason-Jones A. Greater Accra Maternal Services [dataset]. Version Publication. Zenodo; 2026. doi:10.5281/zenodo.22877900

18. Dotse-Gborgbortsi W, Wardrop N, Adewole A, Thomas MLH, Wright J. A cross-sectional ecological analysis of international and sub-national health inequalities in commercial geospatial resource availability. Int J Health Geogr. 2018 May 23;17(1):14. doi:10.1186/s12942-018-0134-z

19. Mpango J, Nabukenya J. A Qualitative Study to Examine Approaches used to Manage Data about Health Facilities and their Challenges: A Case of Uganda. AMIA Annu Symp Proc. 2019;2019:1157–66. PubMed PMID: 32308913; PubMed Central PMCID: PMC7153096.

20. Chikumba PA, Chisakasa G. Towards Geodata Maintenance: A Case of DHIS2 GIS Implementation in Malawi. Journal of Health Informatics in Developing Countries [Internet]. 2018 Sep 22 [cited 2026 Sep 23];12(2). Available from: https://jhidc.org/index.php/jhidc/article/view/176

